# Validation of a decision-analytic model for the cost-effectiveness analysis of a risk-stratified National Breast Screening Programme in the United Kingdom

**DOI:** 10.1101/2022.12.05.22283099

**Authors:** Stuart J Wright, Ewan Gray, Gabriel Rogers, Anna Donten, Katherine Payne

**Affiliations:** Manchester Centre for Health Economics, Division of Population Health, Health Services Research & Primary Care, The University of Manchester, Oxford Road, Manchester, UK

## Abstract

**Background:** This study follows structured frameworks to assess the internal and external validity of a decision-analytic model-based cost-effectiveness of approaches to implement a risk-stratified national breast screening programme (risk-NBSP) in the United Kingdom (UK).

**Methods:** A pre-defined set of steps were used to conduct the process of validation of a published decision-analytic model-based cost-effectiveness analysis of a risk-NBSP (UK healthcare perspective; lifetime horizon; costs (£; 2019). Internal validation was assessed in terms of: descriptive validity; technical validity; face validity. External validation was assessed in terms of: operational validation; convergent validity (or corroboration); predictive validity.

**Results:** The results outline the findings of each step of internal and external validation. The positive aspects of the model in meeting internal validation requirements are shown. The limitations of MANC-RISK-SCREEN are described.

**Conclusion:** Following a transparent and structured validation process, MANC-RISK-SCREEN has been shown to have good internal validity and satisfactory external validity. We suggest that MANC-RISK-SCREEN provides a robust decision-analytic model to assess the cost-effectiveness of risk-NBSP from the UK perspective.

**Key points for decision makers:** There are emerging suggested adaptations to national screening programmes, such as the introduction of risk-stratification to the national breast screening programme (risk-NBSP) in the United Kingdom (UK)

There is a key role for the use of decision-analytic model-based analysis of healthcare interventions, such as a risk-NBSP, that are difficult to evaluate in trials due to the large number of participants required and very long follow up period required.

This study follows structured frameworks to assess the internal and external validity of a decision-analytic model-based cost-effectiveness of a potential risk-NBSP. The decision-analytic model is shown to perform to a satisfactory level, with possible limitations described clearly, to inform resource allocation decisions from the perspective of the UK healthcare system.

## Introduction

Risk-stratified national breast screening programmes (hereafter risk-NBSP) are being suggested as a potential adaptation to existing programmes that offer a mammogram (x-ray of the breast) to all women in a selected age group. Each country, and in some instances regions within a country, each have their own strategy for offering a NBSP. The approach to a NBSP can vary in terms of the; age at which screening is first offered to women in the population (NBSP starting age); interval between screens (NBSP screening interval); age at which screening is stopped (NBSP stopping age); number of x-rays used (one or two-view mammography); supplementary screening technologies used (ultrasound and/or magnetic resonance imaging); interpretation of the x-ray (manual or digital). In the United Kingdom, in 2021, the current NBSP invite women, via a letter sent to their home address, to have a mammogram that is then repeated every three-years. The current eligible age-range for the UK-NBSP starts within three years of reaching their 50^th^ birthday up to the age of 70 years (inclusive) (1).

Factors know to influence a women’s ten-year risk of developing breast cancer have been used to develop risk prediction models developed in various formats, examples include BOEDICEA and the Tyrer-Cuzick survey (2–4). A further necessary requirement has been the development of criteria to categorise woman into specified risk-groups such as low, population risk or high-risk of developing breast cancer in a specified time period (ten-year or life-time). Collectively, the ability to identify a women’s risk of breast cancer, approaches to the feedback of individual risk and actions to suggest given the defined risk-category has stimulated interest and the generation of evidence to support the introduction or risk-NBSP (5–10). To date, there are no examples of a risk-NBSP used in practice but there is consensus about the need for different types of evidence to support their introduction (11).

Generating trial-based clinical evidence of the effectiveness of a risk-NBSP compared with existing approaches to NBSP is neither feasible nor perhaps desirable due to the inherent limitations of the necessary follow-up and the challenges of including women from different risk-group. Within this context, and in keeping with the predominant view, economic evidence to understand the potential value of risk-NBSP should come from decision-analytic model-based analyses using appropriate methods that answer the specified decision-problem (12–15). Tappenden and Chillcott clearly outlined the stages in the development of a decision-analytic model for the purpose of informing resource allocation decision-making (16). An overarching concept described in these stages was the need to include a process in which the decision-analytic model that involves cycles of model checking and validation (16). The concept of iterative economic evaluation mirrors the recommendations made by Sculpher and colleagues. This iterative approach involves building on an early economic analysis to move in cycles, supported by additional data collection, towards a definitive evidence base characterised by a decision-analytic model that decision makers trust and have confidence in the outputs (17).

A suite of recommendations have been developed in the health care context which are designed to enable decision-analysts to have a structured approach to developing, building, and appraising the quality of decision-analytic models (18–23). A crucial element in these recommendations is the need for validation to enable decision-makers using the outputs of a decision-analytic model-based economic evidence to have sufficient confidence in the reported results (24). Enabling decision-makers trust and confidence, by conducting a systematic and transparent process of validation, is a vital component that decision-analysts should take seriously so the decision-analytic model has sufficient credibility (23). A fundamental component supporting the process of validation is the need for transparency in the decision-analytic model structure and use of data. Simplistically, transparency can be achieved by using open source programming languages, such as R, and making the code public (25,26). Using a programming language with the code that can be accessed by anyone is necessary but not sufficient. Decision-analysts have clearly pointed out the need for a structured framework to present code such that it can be read and followed by someone other than the decision-analyst who created the decision-analytic model (27). It then follows logically that the process of validation for a published decision-analytic model should also be transparent.

The concept of validation for decision-analytic models is generally poorly defined (28) and can be conducted using different approaches. The aim of this study was to design and report the validation of a published decision-analytic model designed to conduct a cost-effectiveness analysis of a risk-based NBSP in the UK setting (29). The resulting outputs of this model reporting the healthcare costs and health consequences of a risk-NBSP compared with no screening in the UK setting, will be published separately in a follow-up paper.

## Method

A pre-defined set of steps were used to conduct the process of validation of a published decision-analytic model-based cost-effectiveness analysis (29). There are numerous, inconsistent, definitions of defining the process of decision-analytic model validation (28). This study therefore took a pragmatic approach to describe the required steps of validation that are needed (in a normative sense) to enable the perception of the credibility of a decision-analytic model that was built to address a clearly specified decision problem.

### Description of the original decision-analytic model

The decision-analytic model reported in Gray et al (2015) was developed to address the decision problem: “*What are the key drivers of the incremental costs and benefits of example stratified breast screening programs compared with the current National Breast Cancer Screening Program?*” The key characteristics of the ‘Gray’ decision-analytic model are outlined in Table 1.

**Table 1:** Key characteristics of the original decision-analytic model

| Characteristics | Descriptions |
| --- | --- |
| **Interventions** | Risk-1: a risk-based stratification defined by the Tyrer-Cuzick risk-algorithm enhanced with density and texture measures. Three strata (with associated screening intervals) were defined by ten-year risks of breast cancer of (i) <3.5% (three-yearly); (ii) 3.5 to 8% (two-yearly); (iii) >8% (annual).<br>Risk-2: a risk-based stratification defined by the same algorithm as risk-1 but with strata defined by dividing the population into thirds based on risk (tertiles): (i) the lowest risk tertile (three-yearly); (ii) the middle tertile (two-yearly); (iii) the highest risk tertile (annual).<br>(iii) Masking: current screening approach with supplemental ultrasound offered to women with high breast density, defined using Volpara Density Grades. High risk was defined as greater than an 8% ten-year risk of breast cancer. Women with both high breast density and high risk of breast cancer were offered supplemental MRI instead of ultrasound.<br>Risk-1 with masking: the risk-1 stratification approach together with the strategy described in the masking approach. |
| **Comparators** | Two comparators were defined:<br>Current national breast screening programme (UK-NBSP): Women between 50 and 70 years with screening every three-years using mammography<br>No screening: no use of mammography in the population for screening purposes. All cancers would present with clinical signs or symptoms |
| **Model type** | Discrete event simulation programmed in R |
| **Population** | Women eligible for a national breast screening programme |
| **Setting and perspective** | National healthcare service in the United Kingdom |
| **Time Horizon** | lifetime |
| **Costs** | National currency (£) at 2014 prices |
| **Benefits** | Life-years and quality adjusted life years |
| **Discounting** | 3.5% for both costs and benefits (base case)<br>3.5% for costs and 1.5% for benefits (sensitivity analysis) |
| **Cost-effectiveness threshold** | NICE UK-recommended threshold of £20,000 per QALY gained |

The Gray decision-analytic model was a discrete event simulation comprising three core parts: the stratification (or lack of stratification) of women into different screening intervals depending on their estimated risk of breast cancer, a breast cancer natural history model, and diagnosis and treatment of breast cancers that have been detected by screening. In brief a simulated woman is generated with breast density, 10 year risk of breast cancer, and lifetime risk of breast cancer sampled from a data frame of observations from women who took part in the PROCAS2 study (30). Women are stratified to different intervals of breast cancer screening depending on their ten-year risk of cancer and the screening strategy chosen. Women then attend screening appointments where a possible cancer may be detected depending on the presence of the cancer, its size, and the sensitivity of screening given the size of the tumour. Alternatively, a cancer may be detected clinically between screening appointments. When a tumour occurs, a Nottingham Prognostic Indicator (NPI) grade is assigned with different probabilities depending on the size of the tumour at detection. Utility values, treatment costs, and different survival times are assigned depending on the NPI grade. The total healthcare costs and health consequences (quality-adjusted life-years (QALYs)) are calculated across the cohort of simulated women and can be compared with other screening strategies by re-running the model.

The Gray decision-analytic model was developed in the open source software R, allowing for increased transparency. In its original form, the Gray decision-analytic model was made freely available upon request from the authors in keeping with recommendations from Jansen et al (2019) and Carlsson et al (2019) (31,32). To date the original model has been shared with two academic research teams working in the area of risk-NBSP who have subsequently developed their own models based on this original structure.

A range of clinical and economic inputs are reported, along with their sources, in the published Gray model. Details about the selection of the most appropriate tumour growth model, derivation of breast cancer incidence by age, and a meta-analysis to provide estimates of survival by Nottingham Prognostic Indicator (NPI) grade are provided in the supplementary materials of this original study (29). This supplementary material has subsequently been updated and uploaded onto GitHub to provide full transparency and enable a decision-analysts and person accessing the model to determine if “available input data is appropriate, accurate and sufficient and that data transformations were correctly performed” in line with recommendations provided by AdViSHE (25).

Various clinical and economic outcomes could be generated by the Gray decision-analytic model including but not limited to: average QALYs, average costs, average life years, average number of screens, proportion of women experiencing cancer, and proportion of cancers detected by screening. Deterministic ICERs can be calculated by comparing the estimated mean healthcare costs and QALYs for two or more different screening strategies. Additional R scripts are available to incorporate uncertainty through the conduct and reporting of probabilistic sensitivity analysis (PSA) and a generalised additive model to reduce the computational complexity of the PSA process. The subsequent validated model (hereafter called ‘MANC-RISK-SCREEN’) can also produce these same clinical and outcome outcomes and run deterministic and probabilistic sensitivity analyses.

Due to the large number of parameters in the decision-analytic model and paucity of data with which to fit the model to, calibration of the model was not conducted in the Gray model or MANC-RISK-SCREEN. A key focus was to avoid overfitting the model to the UK national screening context that is the source of the available data for use as input parameters. However, the tumour growth model underlying the economic model has been calibrated to data from the Norwegian breast cancer screening programme (33).

### The components of model validation

In this study, decision-analytic model validation aimed to explore the degree of internal and external validity. Internal validation has been described in terms of three criteria (23,34): descriptive validity to assess whether the degree of simplification used in the decision-analytic model structure still adequately represents the natural history of the specified disease and/or pathways of care; technical validity to assess whether the decision-analytic model was appropriately programmed to produce the intended outputs from the specified inputs; face validity to assess whether the decision-analytic model produces outputs consistent with theoretical basis of disease and the intervention (35). External validation can be described in terms of three criteria: operational validation to assess whether the outputs produced by the decision-analytic model are sufficiently accurate; convergent validity (or corroboration) to compare the decision-analytic model with other published approaches addressing a similar decision-problem; predictive validity to assess whether the outputs produced by the model sufficiently represent outputs from alternative sources.

#### Face validity

Face validity is a type of internal validity that captures first order validation as defined by (36). The process of assessing face validity is intuitive and subjective that requires value judgements to be made by the decision-analyst. These value judgements require the decision-analyst to be explicit about the criteria used when assessing face validity. There is no available published criteria to assess decision-analytic model face validity. Assessing face validity is, therefore, currently reliant on the decision-analyst, with input from relevant clinical expertise, producing an adequate explicit description of if, and how, the outputs are consistent with theoretical basis of disease, the intervention and relevant comparators, for a decision-maker to assess the credibility of the decision-analytic model in this regard.

#### Descriptive validity

Descriptive validity can be viewed as being synonymous with the model conceptualisation process (19). The process of understanding the degree of descriptive validity has also been referred to as conceptual validation as part of published criteria ‘Assessment of the Validation Status of Health-Economic decision models (AdViSHE)’ for assessing model validation to assess: ‘whether the theories and assumptions underlying the conceptual model … are correct and the models representation of the problem entity and the models’ structure, logic and mathematical and causal relationships are ‘reasonable’ for the intended purpose of the model’ (37). Assessing descriptive validity is a subjective process and requires ‘expert’ input from people with relevant knowledge of the disease and intervention being represented by the decision-analytic model and supported by people with relevant technical expertise in decision-analytic modelling. Similar to the application of consensus methods, like Delphi (38), it is also necessary to have a clear threshold what is a ‘sufficient’ level of ‘descriptive validity’ which requires taking account of the purpose of the decision-analytic model (the decision problem).

#### Technical verification

Technical verification is a type of internal validity that captures second order validation (36) and involves a debugging process and assessment of the accuracy of the decision-analytic model in terms of inputs creating ‘valid’ outputs. The process of completing technical verification has been supported by the publication of a verification checklist designed to ‘Reduce Errors in Models and Improve Their Credibility’ called TECH-VER (35). The TECH-VER checklist is a highly detailed list of steps to be used by decision-analysts to reduce the chance of errors in coding the mode structure and calculating data inputs from external data sources (eg. generating measures of overall survival). The TECH-VER checklist does not generate an overall score of technical validity but relies on the decision-analyst describing which criteria are relevant and have been met with a description of how.

#### Operational validation

The process of assessing operational validation is, perhaps, the one most readily interpreted, using lay-terms, as assessing ‘external’ validity. Haji Ali Afzali and colleagues (36) call this third order validation. Operational validation involves comparing decision-analytic model outputs by using different sources of input data that may come from data that was used in the original model (dependent) or data that has been taken from an alternative source (independent) (37). Intuitively, independent operational validation is more robust, in terms of assessing operational validation, than dependent operational validation. However, both independent and dependent validation have key roles when assessing external validity.

#### Predictive validity

The process of assessing predictive validity may not, at first glance, seem relevant to decision-analytic models but rather be the focus for predictive models. Predictive validation is about understanding how well any model predicts future events (36). An alternative interpretation offered by Gray and colleagues is that predictive validity is about seeing the impact on outputs when more data have become available; and in this way it is possible to see if the decision-analytic model has predicted future events (39). The interpretation offered by Gray may, perhaps be better seen as being synonymous with assessing independent operational validation (the approach taken in this study).

#### Cross validation

The process of assessing convergent validity has also been called cross validation. This process requires an alternative decision-analytic model that addresses a similar decision problem to be available. It is most commonly applied for decision-analytic models that have multiplicative purposes rather than in instances when a bespoke structure has been created for a single decision-problem. A well-established process of assessing convergent validity has been set up by the Mount Hood challenge for decision-analytic models in the area of diabetes (24). Convergent validity is also a descriptive process in which the decision-analyst should outline the ways in which different decision-analytic models are the same. Any differences between different decision-analytic models in terms of the outputs produced should be explainable by the decision-analyst to provide assurance of credibility.

### Model validation process

A team of six health economists, supported by an external expert in building decision-analytic models for national decision-making bodies, conducted the model validation process. Each individual had a specific task. The process of model validation involved going through each component part of validation in a step-wise manner. The external expert completed the TECH-VER process. At the end of the model validation process, two published checklists were completed: TECH-VER and AdViSHE. The final MANC-RISK-SCREEN decision-analytic model was then uploaded onto GitHub with all documentation required to understand the model running code, inputs and outputs.

## Results

This section describes the results from the validation of MANC-RISK-SCREEN. The TECH-VER checklists and AdViSHE checklists are reported in Supplementary Appendices 1 and 2, respectively. All code and documentation relating to MANC-RISK-SCREEN are located on GitHub see https://github.com/stuwrighthealthecon/MANC-RISK-SCREEN/tree/main/Model%20Core%20Files/Documentation.

### Validation of MANC-RISK-SCREEN

The development and validation of MANC-RISK-SCREEN built on the original model by Gray and started in February 2021 and together with the independent assessment was completed in June 2022. The process of updating and validating the model took place in discrete steps:

1. Independently reproducing the original Gray decision-analytic model to check for errors and identify areas for improvement;

2. Updating the decision-analytic model to finalise the structure of MANC-RISK-SCREEN;

3. Updating input parameters from the Gray decision-analytic model for MANC-RISK-SCREEN;

4. Checking the face validity of MANC-RISK-SCREEN with stakeholders;

5. Checking the descriptive validity MANC-RISK-SCREEN with stakeholders;

6. Conducting independent technical verification of MANC-RISK-SCREEN;

7. Operational validation of MANC-RISK-SCREEN;

8. Assessing the predictive validity of MANC-RISK-SCREEN for specified targets;

9. Cross validation of MANC-RISK-SCREEN

These steps were performed in sequence. The steps addressed each of the components of the model validation process.

### Reproducing the original model

The decision-analytic model was re-built by a health economist (SJW) not involved in the design and conduct of the original decision-analytic model built by Gray. The health economist (SJW) first read the original R code, including accompanying functions, and wrote a text-based algorithm in Microsoft Word explaining the steps taken in each stage of the model to conduct the analysis. This text-based algorithm was then checked by the lead modeller in the early economic evaluation (EG) who clarified any areas of confusion. This text-based algorithm is available in the documentation area of the GitHub repository for MANC-RISK-SCREEN.

The health economist (SJW) then used the text-based algorithm to reconstruct the model in a new R script and this script was then compared to the original to detect potential errors in both model versions. Only two significant errors (that could influence the estimated cost-effectiveness) were identified during this process. To determine whether a cancer was screen detected a random number was drawn and the cancer determined to be screen detected if the number was greater than a variable representing the proportion of cancers that are screen detected. This should have been the case if the random number was less than the proportion screen detected and was changed in the update model. Fortunately this variable was set to 0.5 and not varied in the PSA so this error had not had an impact on the results. The R code did not include a cost of follow up testing for false-positive screening results thereby potentially overestimating the cost-effectiveness of strategies with more frequent screening. The other changes in the model were made for improvements in the speed of execution, for example defining variables before loops rather than in them, or cosmetic in making the code more readable. An example of the latter was the inclusion of four required R functions in a single accompanying script rather than four individual ones.

### Structural update

When MANC-RISK-SCREEN was built in R structural changes and additional features were included. A key change was that the categorisation of breast tumours was changed from NPI-based classification to a stage-based classification as this significantly increased the availability of relevant data for key input parameters. The start age of the screening was changed to a fixed age of 50 years rather than uniformly varying between 47 and 51 years. This change was made because the varying start age had only been applied to some of the strategies in the early model and was deemed to potentially bias the results. Parameters relating to three types of imperfect uptake for risk-stratified screening were added: uptake for risk prediction; uptake for receipt of risk prediction; uptake for changed screening intervals. A number of additional screening strategies were added to the model including reduced (5 or 6 yearly) screening for women at low risk of breast cancer, and a fully stratified screening programme with more frequent screening for those at higher risk and less frequent screening for those at lower risk.

### Update input parameters

Five years had passed since the publication of the early economic evaluation by Gray in 2017. This time period meant it was likely that values of the parameters used in the Gray decision-analytic model were likely to be out of date. A comprehensive update of decision-analytic model inputs for MANC-RISK-SCREEN was conducted. The process of updating the input parameters is provided in detail in the documentation folder in the model GitHub repository. Systematic reviews were conducted to identify more recent health utility and cost estimates by breast cancer stage. The cost of stratification was updated to incorporate estimates from a microcosting study. New values for screening related parameters were identified from published audits and reports on the status of the NHS breast cancer screening programme. Studies citing the sources of clinical parameters, including the tumour growth model, were searched to determine if any newer appropriate values were available. A full list of final model parameters and distributions for PSA are available in the documentation section of the GitHub repository for MANC-RISK-SCREEN.

### Face validity

Following the reconstruction and parameter update, preliminary results from MANC-RISK-SCREEN were presented at a close-out meeting of main programme of work called PROCAS2 (40). Two suggestions from this meeting were to: include uptake for risk stratification and screening attendance. These two parameters were subsequently added to MANC-RISK-SCREEN and parameterised. Further suggestions to extend the decision-analytic model to estimate the cost-effectiveness of adding single nucleotide polymorphisms (SNPs) to the risk stratification strategy and of adding in the impact of starting women at high risk of breast cancer on preventive medicines are topics for further development of MANC-RISK-SCREEN.

### Descriptive validity

The descriptive validity of the model was assessed on a continual basis by monthly meetings between the six health economists involved in the validation of MANC-RISK-SCREEN. Two of these health economists (KP, SJW) directly interacted with two clinical experts in risk-based breast screening, a statistician involved in generating the risk-prediction model underpinning the Tyrer-Cuzick algorithm and a health psychologist involved in assessing uptake to the PROCAS2 study. The first of these meetings between the health economists set the thresholds for when the model validation process was sufficient. There were discrete thresholds for each component of model validation.

### Technical Verification

To complete technical verification of MANC-RISK-SCREEN involving an assessment of an error-check, an independent experienced R user with expertise in producing decision-analytic models for national decision-making body was approached. This individual was asked to complete a technical verification of a version of the final model using the TECH-VER checklist as a guide. A number of errors were identified in this process. The duration of QALYs experienced was forced to be an integer year and so sometimes patients had higher QALYs than life years. A problem in one of the functions meant that patients diagnosed with cancer sometimes lived longer than they would have done without the cancer. A problem with two of the screening strategies was identified whereby a variable was being called using an out of date name meaning the model would not run. In addition, an error was identified with the use of supplemental screening whereby in some iterations of an if statement no value was assigned to a variable causing problems further on in the model. Other suggestions were made regarding the documentation for the model and potential approaches to speed up analysis time. As at the time of completion no published results of the final model were available, some sections of the checklist could not be completed.

### Operational validation

The operational validation of MANC-RISK-SCREEN involved determining whether the clinical outputs of the model aligned with data on breast cancer epidemiology observed in the UK. Results used in the operational validation come from model output from the scenario of the current (three-year interval) screening programme targeted at women aged between 50 and 70 years.

Operational validation targets were selected based on our belief that close correspondence of these model outputs to targets may increase confidence in model primary cost-effectiveness results (see https://cisnet.cancer.gov/). Target selection was also limited by the availability of target data or summary statistics. Selected targets were related to incidence and detection rates. Survival by cancer stage was also considered as a target but the authors are not aware on any sources of these data for the UK other than those used to generate the input parameters for the model. There were two specific targets for incidence and detection.

- Age-specific cancer incidence under current screening scenario is close to that reported in national cancer incidence statistics. [source: ONS cancer incidence UK 2017 (41)]
- Proportion of breast cancers detected by screening matches the proportion reported in national breast cancer screening audits [source: NHS Digital Official Statistics (42)] and the stage/size distribution of cancers detected at screening and through all diagnostic routes matches that reported in available registry data [Source: CRUK compiled from registries in each nation (43) and NHS Digital Official Statistics (42)].

### Predictive validity

The observed and predicted age-specific breast cancer incidence rates are reported in Table 2 and displayed in Figure 1. The cancer rates were identified to be similar for women before screening age.

**Table 2:** Predicted and Observed Age-specific incidence rates

| Age band (years) | Age-specific breast cancer incidence rates |  |  |  |
| --- | --- | --- | --- | --- |
|  | 2016-2018<br>Observed (per<br>100K)<sup>1</sup> | Predicted using<br>2018-2020 ONS<br>mortality (per<br>100K) | Predicted using<br>2016-2018 ONS<br>mortality (per<br>100K) | % difference |
| 40 to 44 | 124.6 | 113.7 | 113.3 | -8.7 |
| 45 to 49 | 214.8 | 220.7 | 220.9 | 2.8 |
| 50 to 54 | 279.8 | 270.6 | 268.4 | -3.3 |
| 55 to 59 | 285.5 | 275.1 | 273.5 | -3.6 |
| 60 to 64 | 337.9 | 289.3 | 292.9 | -14.4 |
| 65 to 69 | 412.3 | 357.9 | 361.8 | -13.2 |
| 70 to 74 | 372.7 | 348.0 | 354.1 | -6.6 |
| 75 to 79 | 403.0 | 286.9 | 292.1 | -28.8 |
| 80 to 84 | 430.4 | 291.1 | 298.7 | -32.4 |
| 85 to 89 | 447.7 | 296.8 | 300.2 | -33.7 |
| >90 | 448.4 | 307.6 | 268.0 | -31.4 |

<sup>1</sup>Source: Cancer Research UK, 2022 (43)

**Figure 1:**
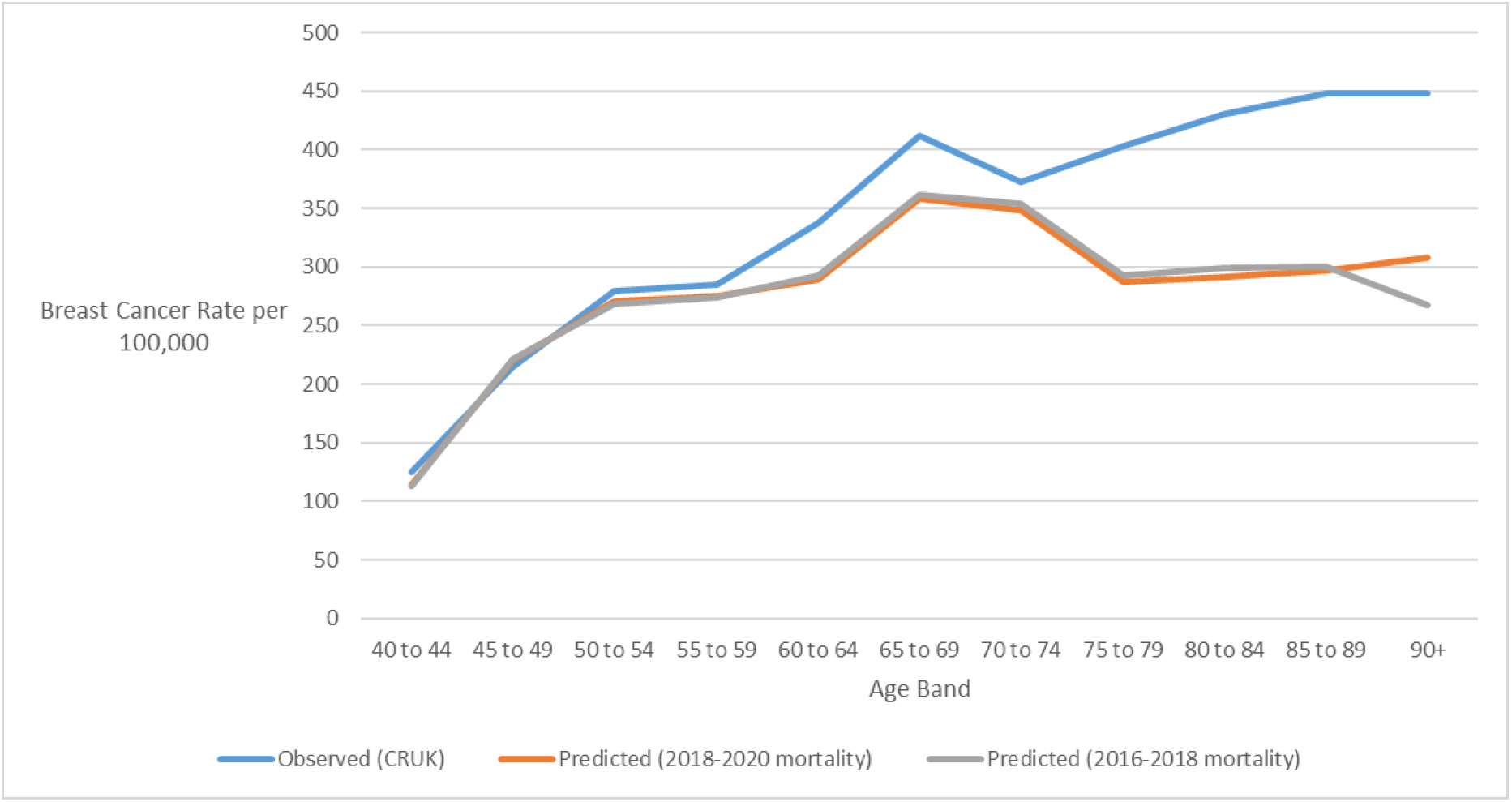
Predicted and Observed Age-specific incidence rate Data source: (43)

After the age of 50 years MANC-RISK-SCREEN appears to underestimate cancer rates compared with the UK registry data from 2016-2018. There was a larger underestimation for the 80+ year age groups however this difference is likely to be unimportant because the incident cancers in these age groups may be little affected by alternative screening programmes and will have a smaller effect on survival outcomes. A potential explanation for this divergence is the use of all-cause mortality data from 2018-2020 which may incorporate higher mortality as age increased due to the beginning of the coronavirus pandemic. The ONS mortality data used to derive life expectancy due to all-cause mortality was subsequently changed to the data that from 2016-2018. This marginally improved the incidence estimates but the pattern of underestimating incidence after the age of 50 years remained.

An alternative explanation for the difference in cancer incidence observed is that there is a difference in the probability a woman will be diagnosed with breast cancer in reality (1 in 7 or 14.3%) compared with the value used in the model (11.8%). The latter lower figure is driven by the average lifetime breast cancer risk for the women who participated in the PROCAS-2 study which is lower than the population average. To determine whether the difference in lifetime breast cancer risk was likely to be the cause of differences in incidence by age, the predicted incidence rates by MANC-RISK-SCREEN were inflated by the proportional difference in lifetime risk (see Figure 2). In this case the model predicted rate appears to more closely track the actual rate more closely, if at a little higher rate. The predicted and observed rates diverge at the age of 70 years, although to a lesser degree than with the unadjusted rates.

**Figure 2:**
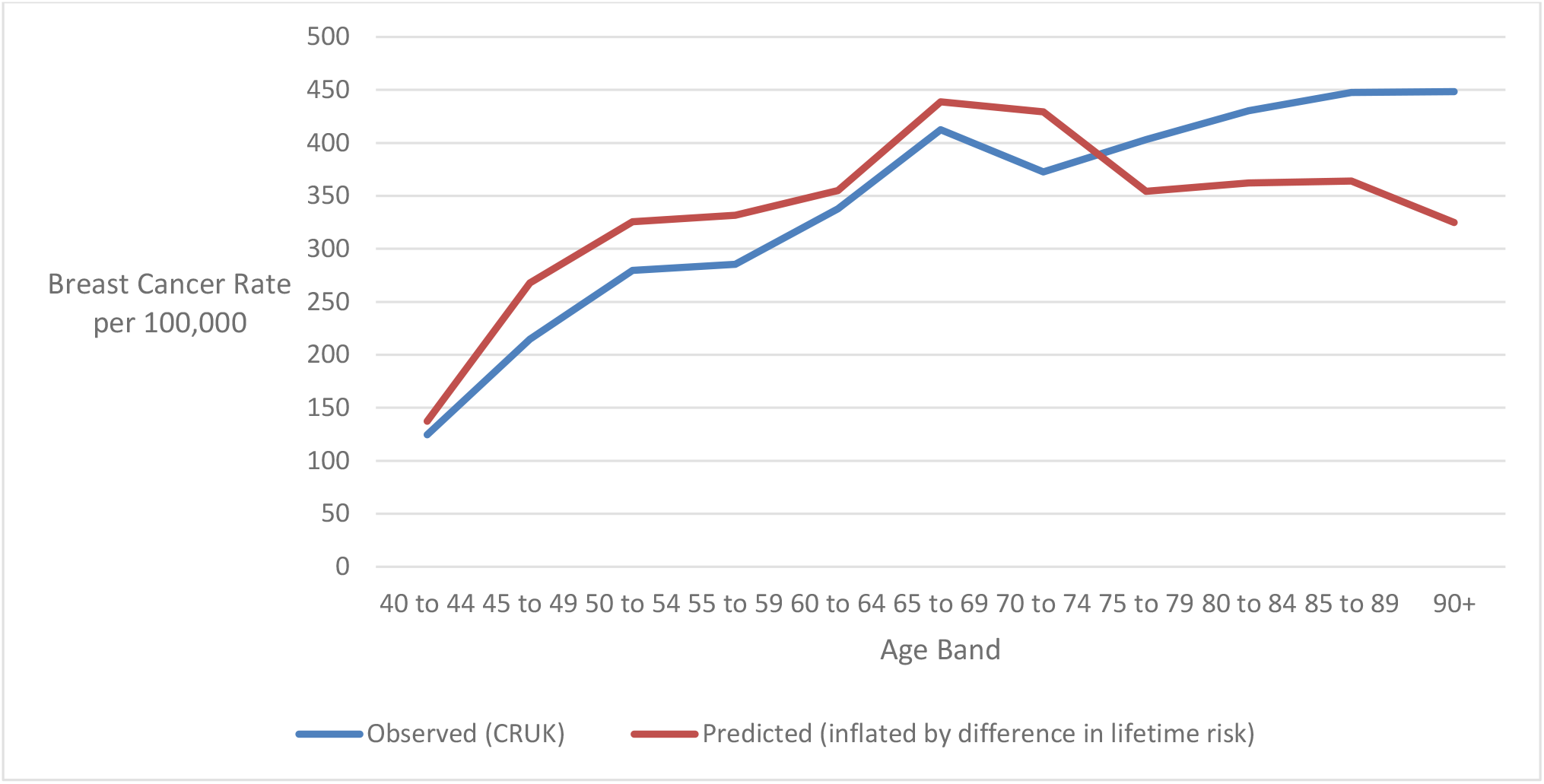
Inflation of predicted incidence rates estimated by MANC-RISK-SCREEN by the proportional difference in lifetime risk Data source: (43)

The most recent breast cancer screening report published by NHS Digital suggested that in the age group eligible for screening, 43% of cancers were identified by screening (42). The original Gray model overestimated the proportion of cancer identified by screening producing a value of 50.2%. A potential explanation for the higher proportion of cancers identified by screening in the Gray model was due to the approach taken to code imperfect screening uptake. In the Gray model, individuals were assigned a probability of 60.5% of attending their first screen. Individuals that had attended at least one screen were assigned an 85.2% probability of attending further screens. In the UK-NBSP it has been observed that women who do not attend their first screen have a reduced likelihood of attending subsequent screens (42). As such, MANC-RISK-SCREEN was recoded such that women had a 60.5% of attending their first screen and if they did not attend this screen only a 19.1% chance of attending subsequent screens. When a woman had attended at least one screen, the probability that she would attend subsequent was increased to 85.2% in MANC-RISK-SCREEN. Following this change, MANC-RISK-SCREEN predicted that 43% of cancers in the age group eligible for screening would be detected by screening and this figure exactly matched the proportion observed in the UK-NBSP.

Table 3 shows the proportions of cancers observed and predicted to be of different stages at diagnosis for cancers diagnosed clinically or by screening. The observed rates are taken form women diagnosed with breast cancer in England (43). Cancers of unknown stage have been excluded from MANC-RISK-SCREEN. The proportion of cancers have been adjusted to incorporate ductal carcinoma in situ that are reported separately. Across all cancers, the Gray model generated too many cancers at stage III (18.9% vs 8% in UK-NBSP) and too few at stage I (28.6% vs 39.4% in UK-NBSP). In addition, the Gray model predicted too few DCIS (5.5% vs 12.9% in UK-NBSP) (44).

**Table 3:** Predicted proportion of clinical and screen detected cancers of different stages

| Stage of Cancer | Observed proportion<sup>1,3</sup> | Gray model predicted proportions | MANC-RISK-SCREEN predicted proportions post validation | MANC-RISK-SCREEN predicted proportions post validation and reversion of cancer by stage matrix |
| --- | --- | --- | --- | --- |
| I | 39.4% | 28.6% | 27.4% | 24.4% |
| II | 35.3% | 41.1% | 39.6% | 39.8% |
| III | 8.0% | 18.9% | 16.1% | 18.8% |
| IV | 4.4% | 5.9% | 6.5% | 6.4% |
| DCIS<sup>2</sup> | 12.9% | 5.5% | 10.5% | 10.6% |

<sup>1</sup>Source Cancer Research UK, 2022 (48)
<sup>2</sup>ductal carcinoma in situ (DCIS)
<sup>3</sup> Proportions adjusted to incorporate DCIS which are reported separately

In the Gray model, it was assumed the DCIS were only diagnosed as part of a UK-NBSP. When a tumour was detected at screening this was assigned to be a DCIS occurrence in 21% of the available cancers regardless of the tumour size. This assumption is likely to be why the proportion of tumours diagnosed as DCIS were considerably lower in the Gray model when compared with the observed estimates in scenarios where DCIS can be diagnosed clinically as well as screen detected. In addition, the approach of allocating any sized cancer as DCIS regardless of size may have affected the stage distribution; DCIS are likely to be smaller than cancers of other stages. In the Gray model the matrix used to determine the probability that a cancer of a given size was of stage I, II or III an assumption had been made using data from source studies (45,46). One of the source studies (Kollias et al, 1999) included estimates in which there was lymph node involvement in a cancer and this was equally likely to involve one or more than one node (45). Cancers with more than one lymph node involved are disproportionately likely to be at a higher stage compared with one or fewer lymph nodes and this may have biased cancers estimated in the Gray model towards a higher stage of diagnosis. To address these issues, MANC-RISK-SCREEN was recoded such that cancer were allocated to a stage or as DCIS based on their size. Data from a study of DCIS was incorporated into the input matrix of the probability of a cancer of a given size being diagnosed at different stages (47). In addition, the proportion of cancers with lymph node involvement in the study where this data was available (Wen et al, 2015) was used to inform the likely level of lymph node involvement in the study where this was not available (Kollias et al, 1999) (45,46).

The predicted proportion of cancers of different stages generated by MANC-RISK-SCREEN is shown in Table 4. The proportion of cancers diagnosed as DCIS closely matches that data from Cancer Research UK (48). The observed values were derived by using the size of cancers diagnosed through the UK-NBSP applying the cancer stage by size matrix (see Github). In some cases, the band of cancer size reported in the UK-NBSP data spanned two bands of cancer size used in the matrix. In these situations, it was assumed that cancer size was evenly distributed across the two bands. Tumours greater than 5cm were assumed to be stage IV at diagnosis. The proportion of DCIS were added from separate data available from Cancer Research UK (48).

**Table 4:** Predicted proportion of screen detected cancers of different stages

| Stage of Cancer | Observed proportion<sup>1</sup> | Gray model predicted proportions | MANC-RISK-SCREEN predicted proportions post validation | MANC-RISK-SCREEN predicted proportions post validation and reversion of cancer by stage matrix |
| --- | --- | --- | --- | --- |
| I | 40.2% | 43.0% | 48.4% | 43.8% |
| II | 23.1% | 17.3% | 17.9% | 19.6% |
| III | 13.1% | 10.8% | 8.3% | 12.3% |
| IV | 2.4% | 3.9% | 5.2% | 5% |
| DCIS | 21.2% | 21.0% | 20.2% | 19.2% |

<sup>1</sup>Based on reported distribution of cancer size at screening. The probability a cancer is of a certain stage given its size is then calculated using combined data from Kollias et al, Wen et al, and Cheng et al (see GitHub)

The distribution of stages for screen detected cancers estimated in MANC-RISK-SCREEN closely matched that observed in the UK-NBSP. However, adjustments subsequently made to the cancer stage by size matrix to improve the fit for the distribution of stages of cancer for all diagnosis routes did not achieve a better match. Therefore the stage by size matrix in the final MANC-RISK-SCREEN model uses a combination of the Wen et al data, which has details of lymph node involvement, and the Kollias et al data with the likelihood of lymph node involvement for different sizes of cancer taken from the Wen et al data.

### Cross validation

There is one alternative decision-analytic model-based economic evaluation of a risk-NBSP published relevant to the UK setting (49). Pashayan et al investigated the cost-effectiveness of alternative risk-NBSP conceptualised as the addition of a risk threshold to the existing age threshold used to determine who is offered screening. On face value, the Pashayan model appears to be directly comparable to the Gray model and MANC-RISK-SCREEN. However, it was not possible to conduct a formal cross validation of the Pashayan model and MANC-RISK-SCREEN in terms of model outputs because the stated decision-problems, decision-analytic model types and structures were not comparable (see Table 5). The Pashayan model evaluated a different risk-based intervention to MANC-RISK-SCREEN. The Pashayan model assumed three-yearly screening would be offered to those above the stated risk threshold. This risk-based intervention was compared with both no screening and the current screening programme (three-yearly screening for all). Each individual’s risk was assessed based on a hypothetical polygenic risk score. A mathematical model was implemented to assess screening effectiveness based on applying a reduction in breast cancer mortality, proportional to breast cancer risk, in a life-table calculation.

**Table 5:**
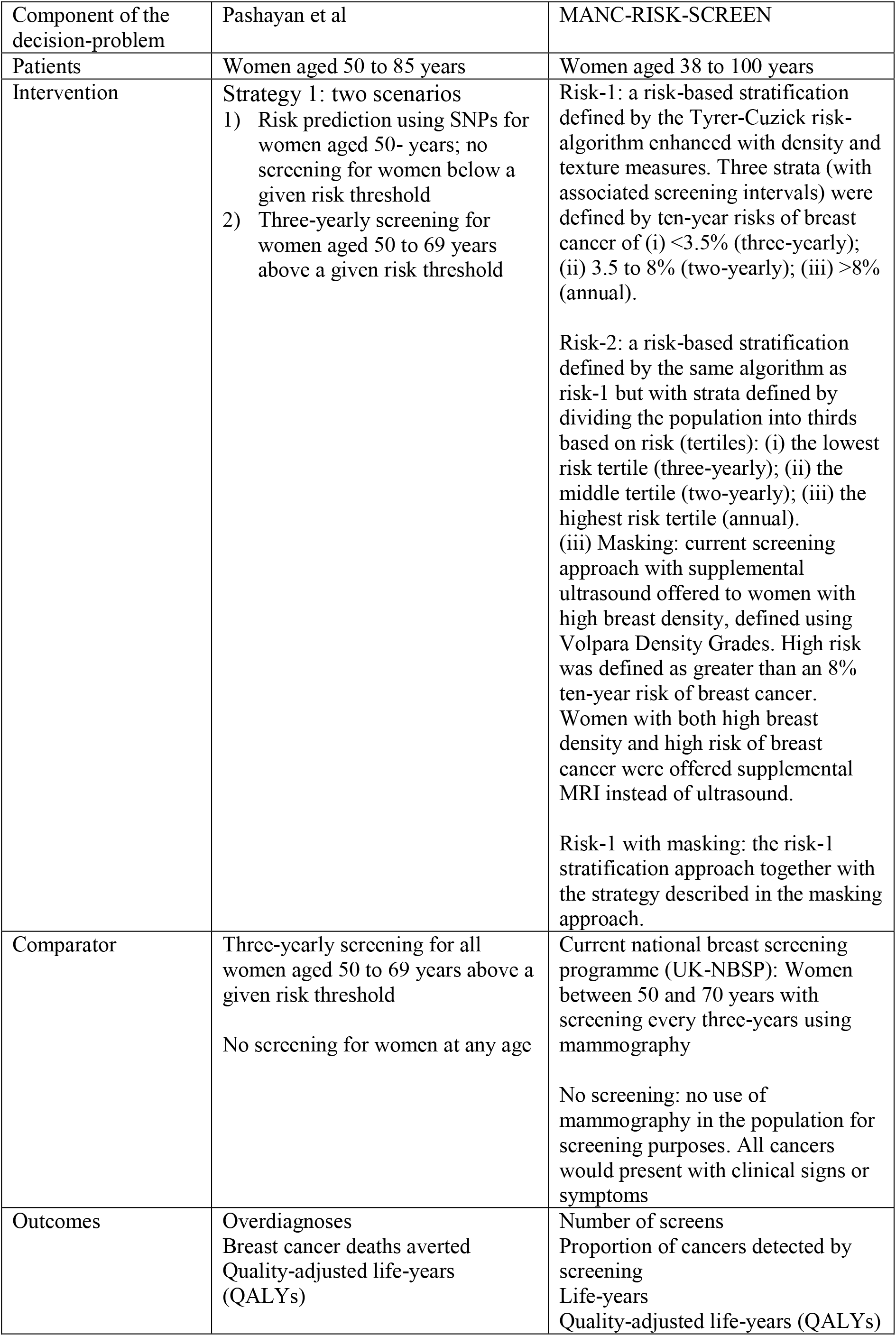
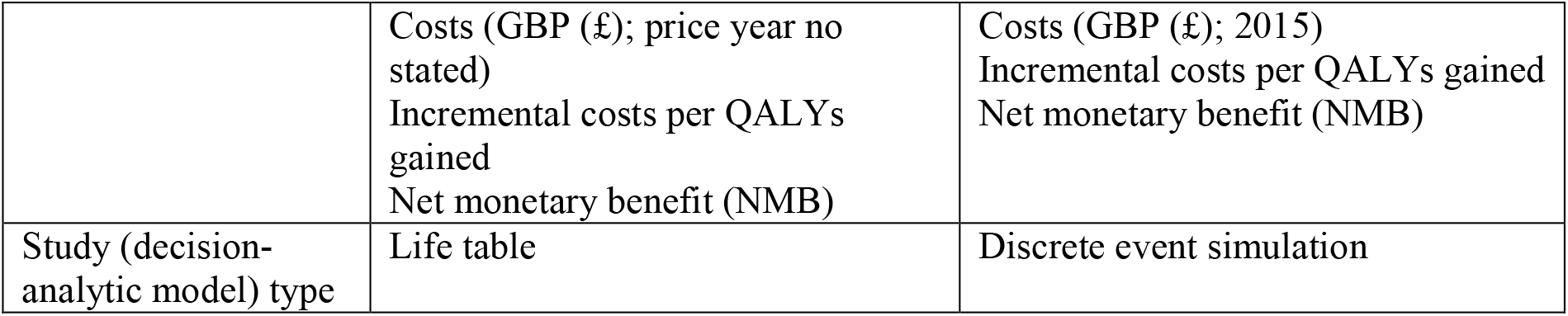
Comparison of the Pashayan model and MANC-RISK-SCREEN

## Discussion

This study reports the validation of a published decision-analytic model-based CEA of a risk-NBSP (29). The Gray decision-analytic model was built to inform an early economic analysis that aimed to suggest the key factors influencing the relative cost-effectiveness of a risk-NBSP rather than produce a definitive economic analysis [29]. We suggest that MANC-RISK-SCREEN has good internal validity that means the decision-analytic model can generate robust estimates of the healthcare costs and health consequences of a risk-NBSP compared with alternative screening strategies. MANC-RISK-SCREEN is now available as an open-source model published on GitHub. A structured and transparent validation process was followed to produce MANC-RISK-SCREEN that is now proposed to be a decision-analytic model with the potential to inform if, and how, healthcare resources should be diverted towards risk-NBSP implemented using different components. The component parts of a risk-NBSP can be varied in terms of the: age at which screening is first offered to women in the population (NBSP starting age); interval between screens (NBSP screening interval); age at which screening is stopped (NBSP stopping age); number of x-rays used (one or two-view mammography); supplementary screening technologies used (ultrasound and/or magnetic resonance imaging); interpretation of the x-ray (manual or digital); approach used to calculate a women’s risk of breast cancer; whether supplementary breast density measurements are taken; classification of the risk categories; approach taken to feedback risk to the women; strategies recommended as a result of identifying a women to be at high risk.

This validation process aimed to maintain a decision-analytic model structure that adequately captures the relevant pathways representing the risk stratification process and subsequent interventions, the current breast screening programme, the natural history of breast cancer and treatment of breast cancer. This aspect of validation was achieved by assessing face and descriptive validity. The input parameters were updated as part of this validation process but this is likely to be an ongoing process, as recommended by Sculpher and colleagues [12], as and when new data become available. The source and steps taken to identify input parameters have been made open source and explicit to facilitate this process of regular updates as required.

The operational validation, together with the assessment of predictive validity, were the main components assessing the external validity of MANC-RISK-SCREEN. This external validation was supplemented by cross validation with a published model. The cross validation was not successful because the only decision-analytic model available for comparison did not match in terms of the interventions used for comparison or model structure. The external validity of MANC-RISK-SCREEN is not perfect. We suggest that decision analysts or decision-makers wanting to use MANC-RISK-SCREEN are aware that it overpredicts clinically diagnosed stage III cancers and underpredicts clinically diagnosed stage I. This overprediciton is likely to affect the results of comparisons of screening programmes compared with no screening.

This paper has built on the recommendations of McCabe and Dixon (13) who proposed a framework for assessing the quality of decision-analytic models such that they are suitable for informing resource allocation decisions in healthcare systems. There are numerous (too many to mention) recommendations and guidelines suggesting the need for, and approaches to, decision-analytic model validation. These have been produced by small groups of individual researchers (for example McCabe and Dixon) or groups of researchers such as the TECH-VER and AdViSHe checklists reaching consensus or task forces as part of international societies such as the International Society for Pharmacoeconomics and Outcomes Research. There are, however, correspondingly few publications that explicitly report the steps completing model validation (50).

### Limitations

The main limitation as part of this validation process was the need to rely on estimating intermediate outcomes generated by the decision-analytic model against data on available from a limited range of sources reporting outcomes. In addition, these data sources only report outcomes from a single scenario; the current screening programme. There is, therefore, limited ability to discriminate between a decision-analytic model that performs well or poorly at the task of predicting comparative cost-effectiveness of alternative screening programmes. The specific targets that were selected for assessing predictive ability were based on the available data sources rather than choosing targets that would be most informative for decision-making.

A related issue to validation of MANC-RISK-SCREEN that needs consideration is the limitations of the risk-prediciton models used to allocate a women’s individual risk. The data sources used to develop these risk-prediciton models means they perform poorly in ethically diverse populations (51). The impact of this limitation will be most apparent if risk-based NBSP are rolled out into practice. In the absence of datasets to assess the predictive value of the risk-prediction models it is impossible to know the extent of the impact of poorly performing risk prediciton on the cost-effectiveness of risk-stratifed NBSP.

## Conclusion

This study has reported a strcutured and transparent validation of a decision-analytic model built to assess the cost-effectiveness of exemplar risk-stratifed NBSP compared with current NBSP or no screening. The validation has suggested MANC-RISK-SCREEN has good internal validity. There are some concerns regarding external validity but these can only be rectified as and when new data sources become available to populate MANC-RISK-SCREEN.

## Data Availability

The R code for the decision-analytic model structure is publicly available in a GitHub repository: https://github.com/stuwrighthealthecon/MANC-RISK-SCREEN

## Acknowledgements

We would like to acknowledge the input of D Gareth Evans, Sue Astley (The University of Manchester), Nico Karssemeijer (Radboud University), Carla van Gils (UMC Utrecht, div. Julius Centrum) into the conceptualisation and structure of the decision-analytic model-based cost-effectiveness analysis that informed the design of this study.

