## Appendices for "Validation of a decision-analytic model for the cost-effectiveness analysis of a risk-stratified National Breast Screening Programme in the United Kingdom"

**Appendix 1:** summary of independent technical verification of MANC-RISK-SCREEN (TECH-VER checklist)

**Appendix 2:** Completion of the Validation-Assessment Tool of Health-Economic Models for Decision Makers and Model (AdViSHE)

### Appendix 1

**Appendix 1:** summary of independent technical verification of MANC-RISK-SCREEN (TECH-VER checklist)

This document provides a detailed description of the technical verification of MANC-RISK-SCREEN conducted by an independent decision-analyst who was not part of the research team that built the original model. The independent decision-analyst followed the general guidance described as part of the TECH-VER checklist[1] to conduct the process of technical verification and produced a written report (included here in this document). The actions the research team took in response to the comments from the independent decision-analyst are also described in this document.

**Overview**
MANC-RISK-SCREEN is an implementation of the decision-model described in: Value in Health: Gray E, Donten A, Karssemeijer N, et al. Evaluation of a Stratified National Breast Screening Program in the United Kingdom: An Early Model-Based Cost-Effectiveness Analysis. Value Health 2017;20:1100–9.

This document describes verification of the model in accordance with the TECH-VER checklist[1]. The model was submitted with all files required to run it, and a brief description of the model along with limited information for model users. A formal user guide was not provided to the reviewer, although it is referenced in the underlying documentation. While some of the model parameters had sources listed as comments in the code, there was no formal write up of the model to compare with, meaning that some aspects of the checklist were not possible. No uncertainty analysis was submitted therefore this section of the TECH-VER checklist was not included.

Despite the lack of a formal user guide, the model code was sufficiently (albeit not generously) commented. This, coupled with the description of the model referenced above enabled verification of the technical structure.

Partway through the review a text algorithm of the underlying functions was provided which enabled a more thorough review of the separate model functions file.

*Subsequent actions taken by the research team*
A text-based algorithm explaining the steps taken in the model is now available in the documentation folder in GitHub. This folder also now contains a list of parameters and their sources and other documents to support users understanding of the model.

#### Running the Model

There was an issue with the code that meant that when **screen_strategy==1** value was set to 5 or 6 the model did not run. This was due to the parameter **start_screening** not being defined in the model code. It was clear from the limited information provided that **start_screening** should have the same value as **screen_startage** and may represent a legacy naming from a previous version of the model.

By setting the value of **start_screening** to **screen_startage** the model ran without issue.

Another issue running the code became apparent when supplemental screening was included. In cases where **VDG>density_cutoff** the if statement did not initialise either **MRI_screening** or **US_screening**. This led to errors further into the model. By initialising these variables inside the if statement where appropriate (and setting them to 0) the model ran without issue.

[1] Büyükkaramikli, N.C., Rutten-van Mölken, M.P.M.H., Severens, J.L. et al. TECH-VER: A Verification Checklist to Reduce Errors in Models and Improve Their Credibility. PharmacoEconomics 37, 1391–1408 (2019). https://doi.org/10.1007/s40273-019-00844-y

*Subsequent actions taken by the research team*
The parameter **start_screening** was a legacy parameter that has since been removed from the model.

Where **start_screening** was mentioned in the model this has now been replaced with **screen_startage** which is the new version of the same parameter.

**MRI_screening** and **US_screening** are were initially defined in the model such that in certain outcomes of an if statement they were not defined but referred to in the model. To rectify this

**MRI_screening** and **US_screening** were initially set to 0 before the if statement was run, solving the issue.

**TECH-VER Checklist**

#### Input Calculations

##### Completeness

The model is self-contained and does not rely on external links of sources to run. The majority of the model is contained within the file MANC_RISK_SCREEN Version 0.R, with population data and a script containing the detailed functions provided separately.

The model is neatly structured with the input parameters listed between lines 70 and 200. Sources are not always specified in the model code.

The model uses inbuilt R libraries to generate values from given distributions, negating the requirement to check formulae line-by-line in many cases.

*Subsequent actions taken by the research team*
A complete list of model input parameters and their sources for the deterministic and probabilistic models is now provided in the documentation folder in the GitHub repository.

##### Black Box Testing

| *Pre-analysis Calculations* | | | |
|---|---|---|---|
| Checklist Question | Results | Comments | *Subsequent actions taken by the research team* |
| Does the technology (drug/device, etc.) acquisition cost increase with higher prices? | Not applicable | Higher screening volumes led to higher costs | Not applicable |
| Does the drug acquisition cost increase for higher weight or body surface area? | Not applicable | | Not applicable |
| Does the probability of an event, derived from an OR/RR/HR and baseline probability, increase with higher OR/RR/HR? | Yes | No relative treatment effects due to structure of the model. Some odds are calculated in the screening_result function and increase with higher input values as excepted | Not applicable |

| | | | |
|---|---|---|---|
| In a partitioned survival model, does the progression-free survival curve or the time on treatment curve cross the overall survival curve? | Not applicable (DES) | The model used functions from R including those from the dqrng package for speed. | Not applicable |
| If survival parametric distributions are used in the extrapolations or time-to-event calculations, can the formulae used for the Weibull (generalized gamma) distribution generate the values obtained from the exponential (Weibull or Gamma) distribution(s) after replacing/transforming some of the parameters? | Not applicable as this model is a DES | | Not applicable |
| Is the HR calculated from Cox proportional hazards model applied on top of the parametric distribution extrapolation found from the survival regression? | Not applicable | | Not applicable |

##### Detailed Comments

No issues were found following this section of the TECH-VER checklist

##### Event/State Calculations

##### Completeness

The number of screens a patient undergoes are calculated in rows 237 – 290.
The patient's time of death is drawn in line 315 from a Weibull distribution defined at row 80. In some cases, the mortality is redrawn if a patient's original mortality was before their cancer mortality (lines 370-380). This is discussed further in the detailed comments below.

The incidence and timing of cancer is calculated from lines 347 to 355 and is based on the underlying risk of a patient (line 232).

The costs are defined in rows 164 to 177 and applied in the DES section of the model (from row 404 onwards).

The utility values by age are set in row 110 with other utility values listed from lines 182 to 207. All QALYs are summed at the end of the DES component (lines 525 onwards.

| Event/State Calculations | | | |
|---|---|---|---|
| Checklist Question | Results | Comments | *Subsequent actions taken by the research team* |
| Calculate the sum of the number of patients at each health state | Not applicable | As this is a DES one patient is simulated on each model loop. | Not applicable |
| Check if all probabilities and number of patients in a state are greater than or equal to 0 | Yes | As this is a DES there are no states. Other probabilistic elements of the model largely use inbuilt R function for calculating probability distributions. No negative probabilities were found. | NOT APPLICABLE |
| Check if all probabilities are smaller than or equal to 1 | Yes | Probabilistic elements of the model largely use inbuilt R functions for calculating probability distributions. No negative probabilities were found. | NOT APPLICABLE |
| Compare the number of dead (or any absorbing state) patients in a period with the number of dead (or any absorbing state) patients in the previous periods? | Not applicable | | NOT APPLICABLE |
| In case of lifetime horizon, check if all patients are dead at the end of the time horizon | Yes | Time horizon can be specified by the user. The line: **if(Mort_age >= time_horizon){Mort_age <- 99.99}---** ensures no patients live past this age | NOT APPLICABLE |
| *Discrete event simulation specific:* Sample one of the 'time to event' types used in the simulation from the specified distribution. Plot the samples and compare the mean and the variance from the sample | Yes | As inbuilt R functions are used this is trivially true. For completeness S=sampling 'clin_detect sample' a million times give the approriate mean and standard deviation | NOT APPLICABLE |
| Set all utilities to 1 | No | LY are determined exactly whereas utilities appear to be applied per year. Thus the model predicts slightly more QALYs than life years when the discount rate is set to 0 and all utilities are set to 1. This would be easy to correct by adjusting the final QALY value by the proportion of | Code was added to the model to ensure that utility values could be applied for parts of years. This was achieved by weighting the utility values for specific years in |

| | | | |
|---|---|---|---|
| | | the year that was survived. The line **if(QALY_length<1){QALY _length <-1}** makes it appear as though this choice (that patients should not die mid-year) is deliberate although any assumption to this effect should be stated explicitly. | the QALY vector by the proportion of the year a person spent in different health states |
| Set all utilities to 0 | Yes | Utilities are in two separate sections of the inputs, one for the QoL by age and one for decrements based on health states. For clarity these two sections could be placed next to each other | The specified lines of code were moved next to each other in the script |
| Decrease all state utilities simultaneously (but keep event-based utility decrements constant) | Yes | Halving annual utility reduces in a value approximately half (within Monte Carlo error tolerance) | NOT APPLICABLE |
| Set all costs to 0 | Yes | No costs accrued in model | NOT APPLICABLE |
| Put mortality rates to 0 | Yes | If background death age and cancer death age set time horizon LY = length of model for all patients | NOT APPLICABLE |
| Put mortality rate at extremely high | Yes | It is notable that the mortality rate is redrawn if a patient experiences cancer, this means that two mortality rates need to be set to ensure patients die immediately. | NOT APPLICABLE |
| Set the effectiveness-, utility-, and safety-related model inputs for all treatment options equal | Yes | The screening strategy details are set between lines 235 and 288. By setting all strategies to be equal at this stage the results trivially become equal for all options | NOT APPLICABLE |
| In addition to the inputs above, set cost-related model inputs for all treatment options equal | Yes | The screening strategy details are set between lines 235 and 288. By setting all strategies to be equal at this stage the results trivially become equal for all options | NOT APPLICABLE |
| Change around the effectiveness-, utility- and safety-related model inputs between two treatment options | Yes | The screening strategy details are set between lines 235 and 288. By setting all strategies to be equal at this | NOT APPLICABLE |

| | | | |
|---|---|---|---|
| | | stage the results trivially become equal for all options | |
| Check if the number of alive patients estimated at any cycle is in line with general population life-table statistics | Yes | Model uses Weibull distribution where a=8.97 and b = 86.74. the resulting distribution appears plausible | NOT APPLICABLE |
| Check if the QALY estimate at any cycle is in line with general population utility estimates | Yes | Utility values by age are set in line 111, no source is given however the utilities lower as age increases which is expected behaviour and all values appear reasonable given UK average QALY values | The input parameter sources are now provided in the documentation folder |
| Set the inflation rate for the previous year higher | Not applicable | No specific mechanism for adding a rate of inflation. The costs are listed without sources. Increasing the costs increases total cost output as expected | NOT APPLICABLE |
| Calculate the sum of all ingoing and outgoing transition probabilities of a state in a given cycle | Not applicable | | NOT APPLICABLE |
| Calculate the number of patients entering and leaving a tunnel state throughout the time horizon | Not applicable | | NOT APPLICABLE |
| Check if the time conversions for probabilities were conducted correctly. | Not applicable | | NOT APPLICABLE |
| *Decision tree specific:* Calculate the sum of the expected probabilities of the terminal nodes | Not applicable | | NOT APPLICABLE |
| *Patient-level model specific:* Check if common random numbers are maintained for sampling for the treatment arms | No | Whilst R offers the ability to fix streams of random numbers with set.seed this model relies on large number of randomly drawn patients to be modelled to reduce Monte Carlo error to acceptably small values which negates the requirement to fix streams of | Speed improvements including variance reduction techniques like that described are planned for inclusion in future model version |

| | | | |
|---|---|---|---|
| | | random numbers. Despite this there is no reason that random streams could not be fixed to more closely align each model run if desired. | |
| *Patient-level model specific:* Check if correlation in patient characteristics is taken into account when determining starting population | Yes | The risk matrix that is sampled is generated from applying the R synthpop package to real Volpara breast density estimates from over 150,000 women. Synthpop will retain the original structure of the data (including correlations) | NOT APPLICABLE |
| Increase the treatment acquisition cost | Yes | Costs are not reported by year, however increasing the screening cost increases the overall cost (where screening is carried out) | NOT APPLICABLE |
| *Population model specific:* Set the mortality and incidence rates to 0 | NOT APPLICABLE | | NOT APPLICABLE |

#### Detailed Comments

The checklist above revealed a disconnect between costs and QALYs (when all discount rates are set to 0 and QoL =1 for all states). A review of the relevant section of the code revealed that there is a specific line which sets the length of life for a given year to 1, even if this would exceed the total life years predicted by the model. This should either be listed as an assumption of corrected.

In some cases, the mortality is redrawn if a patient's original mortality was before their cancer mortality (lines 370-380). If the original Weibull distribution mortality is drawn from is accurate then this step may cause issues with the model calculated mortality distribution. In practice, this issue occurs only in rare cases and is unlikely to impact the model materially.

The user guide to the model suggests running many million simulations to minimise Monte Carlo error which should negate any requirement to draw the sample of patients for each screening option. Drawing the same random stream of patients would seem logical, given that there are only 15613 risk profiles to choose from, it may make more sense to the reviewer to sample each patient a given number of times. When combined with the R function set.seed() identical random draws could be applied to each treatment arm. See Appendix 1 for more details.

*Subsequent actions taken by the research team*

Code was added to the model to ensure that utility values could be applied for parts of years. This was achieved by weighting the utility values for specific years in the QALY vector by the proportion of the year a person spent in different health states

Given its likely rarity no action was taken about the issue highlighted with the mortality distribution. Speed improvements including variance reduction techniques are planned for inclusion in future model version. For example, some calculations such as putting an individual in a VDG group could be applied to the synthetic sample rather than being done for each run in the simulation.

#### Additional checks on functions

*Incidence_function*

This function calculates the incidence of cancer and detection. The risk matrix that the values are drawn from contains two cancer time-based risk values, a ten-year risk (TYR), and a lifetime risk. The lifetime risk is used to calculate the likelihood of cancer occurring and the TYR is used (not in this function) to assign a screening risk group.

When the cancer incidence time is calculated the TYR value is ignored, and the time is sampled from the included ONS dataset. The TYR calculated by the model is not included in the calculation and the TYR value does not correspond to the ten-year risk for any simulated patient (with the model start_age set to 38, given that cancer is present there is around an 11% chance of the cancer occurring in the first ten years, whereas the risk data suggests that 24% of all cancers occur in the first 10 years {TYR/Lifetime Risk}).

Furthermore, the risk matrix suggests that the likelihood of a cancer occurring within the first ten years given that cancer occurs varies between women, whereas the current model structure does not have the ability to vary this conditional probability between patients.
In summary, the model uses TYR to stratify screening strategies, but then generates TYR values that do not match.

*Subsequent actions taken by the research team*

We believe this problem arises because of a non-linear relationship between predicted 10 year risk and lifetime risk in the Tyrer-Cuzick algorithm. At higher lifetime risks of cancer, the 10 year risk is disproportionately higher suggesting that women at higher risk of cancer are more likely to have cancer at an earlier age. In the model, the probability of a cancer occurring in the next 10 years is effectively fixed given the distribution of cancers at different ages across the population. As a result, the model may underestimate the number of cancers occurring in high risk women at younger ages.

As few women have sufficiently high 10 year risk, there is likely to be relatively little effect on the outcomes of the model. However, the effectiveness of increased screening in high risk groups may be marginally underestimated. Attempts will be made in future versions of the model to resolve this issue.

*NPI_by_size*

This function uses the tumour size and screen detection status to determine the NPI category using parameters defined in the model initialisation. No issues were found in the implementation and it appears to perform as expected.
*screening_result*

This function calculates the likelihood of cancer being detected by various screening options.
In the line MRI_supp_odds <- ((Sensitivity*6)/(1-Sensitivity))*((MRI_cdr+Mammo_cdr)/Mammo_cdr) it is not clear why the initial sensitivity is multiplied by 6, particularly as US_supp_odds does not contain a value in this location.

*Subsequent actions taken by the research team*

This appears to have been a typing error and the *6 has been removed

*Ca_survival_time*

This function calculates the expected mortality age if cancer is experienced.

The model behaves as described in the text algorithm guide supplied, with distributions values are drawn from producing realistic survival times, with appropriate limits set according to the modelled time horizon.

In some (rare) cases this function results in patients who experience cancer surviving for longer than they would have survived without cancer. The appropriateness of this should be reviewed.

*Subsequent actions taken by the research team*
A line of code has been added which that states that if someone is predicted to live longer than their all-cause mortality date of death with cancer then they die at their all-cause mortality date of death not cancer date of death

#### Results Calculations

##### Completeness

The model requires the user to select a screening strategy and outputs patient level as well as aggregated. Results are presented in absolute terms and the user is required to calculate incremental results outside the provided model.

No write-up including results was submitted for review therefore the TECH-VER checklist was followed as closely as possible, generating results where required to answer the checklist as required. These could not then be compared to any existing results.

The checklist included many questions where the model was directionally correct (more screening, higher costs) however this level of verification will always be weaker than comparing to exact model outputs.

The lack of pre-built results module to calculate and present incremental results may hinder the model's wider appeal and it is recommended that this is added. In the context of the existing complex model, this would be a relatively minor addition and would improve the user experience.

*Subsequent actions taken by the research team*
A pre-built results module and user interface will be added in a future version of the model

##### Black Box Testing

| Result Calculations | | | |
|---|---|---|---|
| Checklist Question | Result | Comments | Actions Taken |
| Check the incremental life-years and QALYs gained results. Are they in line with the comparative clinical effectiveness evidence of the treatments involved? | Yes | Higher screening strategies are more expensive and detect more cancer (higher QALY values) | NOT APPLICABLE |
| Check the incremental cost results. Are they in line with the treatment costs? | Yes | Taking screening to be the treatment, this is unknown as costs reported as a whole. A line by line review of the costs section of the code (line 406 onwards) shows that discount rates are correctly applied. It appears that screening costs are assumed to happen at the | A half-cycle correction has now been applied to costs |

| | | | |
|---|---|---|---|
| | | start of each year, if not listed an assumption then it may be possible to apply a half cycle correction. | |
| Total life years greater than the total QALYs | Yes | | NOT APPLICABLE |
| Undiscounted results greater than the discounted results | Yes | Undiscounted costs not reported however running the model with a discount rate of 0 verifies that this is correct | NOT APPLICABLE |
| Divide undiscounted total QALYs by undiscounted life years | See comment in previous section regarding QALYs and Lys | Undiscounted QALYs not reported however running the model with a discount rate of 0 verifies that this is correct | NOT APPLICABLE |
| Subgroup analysis results: How do the outcomes change if the characteristics of the baseline change? | Not applicable | No subgroup analysis applied, although the model stratifies patients into risk cohorts | NOT APPLICABLE |
| Could you generate all the results in the report from the model (including the uncertainty analysis results)? | Not applicable | | NOT APPLICABLE |
| Do the total life-years, QALYs, and costs decrease if a shorter time horizon is selected? | Yes | | NOT APPLICABLE |
| Is the reporting and contextualization of the incremental results correct? | Not applicable | | NOT APPLICABLE |
| Are the reported ICERs in the fully incremental analysis non-decreasing? | Not applicable | | NOT APPLICABLE |
| If disentangled results are presented, do they sum up to the total results (e.g. different cost types sum up to the total costs estimate)? | Not applicable | | NOT APPLICABLE |
| Check if half-cycle correction is implemented correctly (total life-years with | No | No half-cycle correction applied. As this is a DES there is no | A half-cycle correction has now been applied to costs and outcomes. |

| | | | |
|---|---|---|---|
| half-cycle correction should be lower than without) | | ambiguity about number of patients surviving to time X. however it could be argued that if discounting is applied continuously (as opposed to in annual steps) a half-cycle should be applied to the total QALYs accrued at the end (e.g a patient with QoL=1 for one year from time 0 does not accumulate 1 QALY if the discount rate is applied continuously). Possible that the utilised QALY values already take this into account however as no source is given it is not possible to tell. | |
| Check the discounted value of costs/QALYs after 2 years | Yes | Undiscounted values not reported however running the model with a discount rate of 0 verifies that this is correct | NOT APPLICABLE |
| Set discount rates to 0 | The discounted and undiscounted results are the same | Not reported however running the model with discount rate = 0 verifies this. | NOT APPLICABLE |
| Set mortality rate to 0. Are the total life-years per patient should be equal to the length of the time horizon | Yes, if integer values selected for LY (see previous comments regarding LY) | | NOT APPLICABLE |
| Put the consequence of adverse event/discontinuation to 0 (0 costs and 0 mortality/utility decrements) | Not applicable | | NOT APPLICABLE |

| | | | |
|---|---|---|---|
| Divide total undiscounted treatment acquisition costs by the average duration on treatment | Not applicable | | NOT APPLICABLE |
| Set discount rates to a higher value | Not applicable | | NOT APPLICABLE |
| Set discount rates of costs/effects to an extremely high value | Model performs as expected | Manual code review of the cost/ QALY sections verifies that discounting is applied correctly | NOT APPLICABLE |
| Put adverse event/discontinuation rates to 0 and then to an extremely high level | Not applicable | | NOT APPLICABLE |
| Double the difference in efficacy and safety between the new intervention and comparator, and report the incremental results | Not applicable | | NOT APPLICABLE |
| Do the same for a scenario in which the difference in efficacy and safety is halved | Not applicable | | NOT APPLICABLE |

#### Further Comments

While there are some minor questions regarding the implementation of half-cycle corrections/ discounting the results seemed in line with expected costs/ QALYs given. It is extremely unlikely that the points raised above will materially impact the model in any way.

**Conclusion**

The model passed the vast majority of the TECH-VER checklist questions (where sufficient information was submitted to allow the question to be answered). The main challenge while reviewing the model was that in its current state it falls short of a complete written-up model report for which the TECH-VER checklist is designed. Not only did this mean that some of the sections could not be answered at all, it introduced some ambiguity for other sections, where the questions were answered on a best endeavours basis. Whilst this is useful it is likely to be less rigorous than checks carried out against a model with thorough documentation, pre-calculated results and a formal write-up.

Despite the above, the technical sections of the checklist were mostly investigable. With background information limited to the existing model used as a base for this implementation, a short model overview and comments within the code it is a credit to the model development team that a model of this complexity could be followed with such limited formal documentation.

Although there were issues found in the model review, the issues with the black box tests were minor and unlikely to materially impact the model. It is plausible that most or all of these issues were a result of underlying assumptions which were not submitted for review.

A detailed review of the underlying functions also raised queries about cancer risk profiles for individual patients covering inconsistencies regarding lifetime risk, ten-year risk and mortality which should be reviewed for appropriateness by the development team.

### Appendix 2

**Appendix 2:** Completion of the Validation-Assessment Tool of Health-Economic Models for Decision Makers and Model (AdViSHE)

**Background**

AdViSHE contains 13 items that modellers can complete to report on the efforts performed to improve the validation status of their health-economic (HE) decision model. The tool is not intended to replace validation by model users but rather to inform the direction of validation efforts and to provide a baseline for replication of the results. In addition to using it after a model is finished, AdViSHE can be used to guide validation efforts during the modelling process. The modellers are asked to comment on the validation efforts performed while building the underlying HE decision model and afterwards. Many of the items can be answered simply by referring to the model documentation.

AdViSHE is divided into five parts, each covering an aspect of validation:

- Part A: Validation of the conceptual model (2 questions)
- Part B: Input data validation (2 questions)
- Part C: Validation of the computerized model (4 questions)
- Part D: Operational validation (4 questions)
- Part E: Other validation techniques (1 question)

No final validation score is calculated, as the assessment of the answers and the overall validation effort is left to the model users. It is assumed that the model has been built according to prevailing modelling and reporting guidelines. For instance, the model builders would presumably adhere to the ISPOR-SMDM1 Modeling Good Research Practices (Caro et al., 2010) and/or CHEERS Statement (Husereau et al., 2013). Some questions may not be applicable to a particular model. If this is the case, the model builder should take the opt-out option and provide a justification of why this item is not deemed applicable.

**Validation of the conceptual model (2 questions)**

Part A discusses techniques for validating the conceptual model. A conceptual model describes the underlying system (e.g., progression of disease) using a mathematical, logical, verbal, or graphical representation.

| |
|---|
| *Please indicate where the conceptual model and its underlying assumptions are described and justified*<br><br>'Description of the original model' in main text<br>'Model Conceptualization and Structure' in original publication, to which readers are appropriately directed<br>GitHub repository, to which readers are appropriately directed |
| Face validity testing (conceptual model)<br>***Have experts been asked to judge the appropriateness of the conceptual model?***<br>*If yes, please provide information on the following aspects:*<br>*- Who are these experts?*<br>*- What is your justification for considering them experts?*<br>*- To what extent do they agree that the conceptual model is appropriate?*<br>*If no, please indicate why not.*[2]<br><br>Yes. The original publication, to which readers are appropriately directed, notes that the 'model structure was conceptualized… with input from key clinical members in the ASSURE team (n = 5) and external experts (n = 15).'<br>Little detail of experts' credentials. It is implicit that the experts considered the model appropriate, but no detail on how they influenced its development. |
| Cross validity testing (conceptual model)<br>***Has this model been compared to other conceptual models found in the literature or clinical textbooks?***<br>*If yes, please indicate where this comparison is reported.*<br>*If no, please indicate why not.*<br><br>The original publication notes that the conceptualisation followed an earlier systematic review of published models (Evans et al. 2016), which identified 'no relevant existing models that could be used without extensive modification'. |

[2] Aspects to judge include: appropriateness to represent the underlying clinical process/disease (disease stages, physiological processes, etc.); and appropriateness for economic evaluation (comparators, perspective, costs covered, etc.).

**Input data validation (2 questions)**
Part B discusses techniques to validate the data serving as input in the model. These techniques are applicable to all types of models commonly used in HE modelling.

| |
|---|
| *Please indicate where the description and justification of the following aspects are given:*<br>- *search strategy;*<br>- *data sources, including descriptive statistics;*<br>- *reasons for inclusion of these data sources;*<br>- *reasons for exclusion of other available data sources;*<br>- *assumptions that have been made to assign values to parameters for which no data was available;*<br>- *distributions and parameters to represent uncertainty;*<br>- *data adjustments: mathematical transformations (e.g., logarithms, squares); treatment of outliers; treatment of missing data; data synthesis (indirect treatment comparison, network meta-analysis); calibration; etc*<br><br>'Update parameters' in main text and appendix @@<br>'Model Input Parameters' in original publication, to which readers are appropriately directed<br>GitHub repository, to which readers are appropriately directed |
| Face validity testing (input data)<br>***Have experts been asked to judge the appropriateness of the input data?***<br>*If yes, please provide information on the following aspects:*<br>*- Who are these experts?*<br>*- What is your justification for considering them experts?*<br>*- To what extent do they agree that appropriate data have been used?*<br>*If no, please indicate why not.*[3]<br><br>Experts advised on some parameters for which published data could not be identified (see 'Model Input Parameters' in original publication). No explicit report that experts provided face validation of parameters drawn from published literature. |
| Model fit testing<br>***When input parameters are based on regression models, have statistical tests been performed?***<br>*If yes, please indicate where the description, the justification and the outcomes of these tests are reported.*<br>*If no, please indicate why not.*[4]<br><br>No. Tumour growth model was calibrated to match Norwegian observational data from before and after the start of a population-based screening programme, but no details of adequacy of fit reported. |

3 Aspects to judge may include but are not limited to: potential for bias; generalizability to the target population; availability of alternative data sources; any adjustments made to the data.
4 Examples of regression models include but are not limited to: disease progression based on survival curves; risk profiles using regression analysis on a cohort; local cost estimates based on multi-level models; meta regression; quality-of-life weights estimated using discrete choice analysis; mapping of disease-specific quality of-life weights to utility values.
Examples of tests include but are not limited to: comparing model fit parameters ($R^2$, Akaike information criterion (AIC), Bayesian information criterion (BIC)); comparing alternative model specifications (covariates, distributional assumptions); comparing alternative distributions for survival curves (Weibull, lognormal, logit); testing the numerical stability of the outcomes (sufficient number of iterations); testing the convergence of the regression model; visually testing model fit and/or regression residuals.

**Validation of the computerized model (4 questions)**
Part C discusses various techniques for validating the model as it is implemented in a software program.

| |
|---|
| *If there are any differences between the conceptual model (Part A) and the final computerized model, please indicate where these differences are reported and justified.*<br><br>Not reported |
| External review:<br>***Has the computerized model been examined by modelling experts?***<br>*If yes, please provide information on the following aspects:*<br>*- Who are these experts?*<br>*- What is your justification for considering them experts?*<br>*- Can these experts be qualified as independent?*<br>*- Please indicate where the results of this review are reported, including a discussion of any unresolved issues.*<br>*If no, please indicate why not.*[5]<br><br>Yes. The external expert is experienced at developing decision-models using the same software used for this study (e.g. for NICE). His independence is assured as he works at a different institution. Results are reproduced in appendix @@. The paper details the impact of this step on model development in 'Technical Verification'. |
| Extreme value testing<br>***Has the model been run for specific, extreme sets of parameter values in order to detect any coding errors?***<br>*If yes, please indicate where these tests and their outcomes are reported.*<br>*If no, please indicate why not.*[6]<br><br>Yes; see TECH-VER checklist in appendix @@. |
| Testing of traces<br>***Have patients been tracked through the model to determine whether its logic is correct?***<br>*If yes, please indicate where these tests and their outcomes are reported.*<br>*If no, please indicate why not.*[7]<br><br>Yes; see TECH-VER checklist in appendix @@. |

5 Aspects to judge may include but are not limited to: absence of apparent bugs; logical code structure optimized for speed and accuracy; appropriate translation of the conceptual model.

6 Examples include but are not limited to: zero and extremely high (background) mortality; extremely beneficial, extremely detrimental, or no treatment effect; zero or extremely high treatment or healthcare costs.

7 In cohort models, this would involve listing the number of patients in each disease stage at one, several, or all time points (e.g., Markov traces). In individual patient simulation models, this would involve following several patients throughout their natural disease progression.

| |
|---|
| Unit testing<br>***Have individual sub-modules of the computerized model been tested?***<br>*If yes, please provide information on the following aspects:*<br>*- Was a protocol that describes the tests, criteria, and acceptance norms defined beforehand?*<br>*- Please indicate where these tests and their outcomes are reported.*<br>*If no, please indicate why not.*[8]<br><br>Not formally; however, the TECH-VER checklist in Appendix 1 details how model scrutiny involved testing of individual functions within the model code. |

**Operational validation (4 questions)**
Part D discusses techniques used to validate the model outcomes.

| |
|---|
| Face validity testing (model outcomes)<br>***Have experts been asked to judge the appropriateness of the model outcomes?***<br>*If yes, please provide information on the following aspects:*<br>*- Who are these experts? - What is your justification for considering them experts?*<br>*- To what extent did they conclude that the model outcomes are reasonable?*<br>*If no, please indicate why not.*[9]<br><br>Not reported. |
| Cross validation testing (model outcomes)<br>***Have the model outcomes been compared to the outcomes of other models that address similar problems?***<br>*If yes, please provide information on the following aspects:*<br>*- Are these comparisons based on published outcomes only, or did you have access to the alternative model?*<br>*- Can the differences in outcomes between your model and other models be explained?*<br>*- Please indicate where this comparison is reported, including a discussion of the comparability with your model.*<br>*If no, please indicate why not.*[10]<br><br>The original publication briefly compares outputs (ICERs) with previous publications, but does not compare clinical outputs or explore reasons for discrepancies. |
| Validation against outcomes using alternative input data<br>***Have the model outcomes been compared to the outcomes obtained when using alternative input data?***<br>*If yes, please indicate where these tests and their outcomes are reported.*<br>*If no, please indicate why not.*[11]<br><br>No. This is not possible due to a lack of possible data. |

[8] Examples include but are not limited to: turning sub-modules of the program on and off; altering global parameters; testing messages (e.g., warning against illegal or illogical inputs), drop-down menus, named areas, switches, labelling, formulas and macros; removing redundant elements.
[9] Outcomes may include but are not limited to: (quality-adjusted) life years; deaths; hospitalizations; total costs.
[10] Other models may include models that describe the same disease, the same intervention, and/or the same population.
[11] Alternative input data can be obtained by using different literature sources or datasets, but can also be constructed by splitting the original data set in two parts, and using one part to calculate the model outcomes and the other part to validate against.

Validation against empirical data

***Have the model outcomes been compared to empirical data?***

*If yes, please provide information on the following aspects:*

*- Are these comparisons based on summary statistics, or patient-level datasets?*

*- Have you been able to explain any difference between the model outcomes and empirical data?*

*- Please indicate where this comparison is reported.*

*If no, please indicate why not.*

Comparison against the data sources on which the model is based (dependent validation)

Not reported.

Comparison against a data source that was not used to build the model (independent validation)

Extensive validation of this type reported in 'Independent Operational Validation', comparing model outputs with multiple national datasets:

- age-specific cancer incidence (compared with ONS cancer statistics)
- proportion of cancers detected by screening (compared with NHS Digital Official Statistics)
- stage and size distribution of cancers detected at screening and through all diagnostic routes (compared with CRUK data and NHS Digital Official Statistics).

Differences between model and empirical data are discussed (partially due to dimensions in which PROCAS participants differ from the wider population from whom the national statistics are drawn).

**Other validation techniques (1 question)**

Other validation techniques

*Have any other validation techniques been performed?*

*If yes, indicate where the application and outcomes are reported, or else provide a short summary here.*[12]

Replication of original model with new code (reported in 'Reproducing the original model') is a form of double-programming that should give confidence to the robustness of both implementations.

Source: Vemer, P., Corro Ramos, I., van Voorn, G.A.K. *et al.* AdViSHE: A Validation-Assessment Tool of Health-Economic Models for Decision Makers and Model Users. *PharmacoEconomics* **34,** 349–361 (2016). https://doi.org/10.1007/s40273-015-0327-2

[12] Examples of other validation techniques: structured "walk-throughs" (guiding others through the conceptual model or computerized program step-by-step); naïve benchmarking ("back-of-the-envelope" calculations); heterogeneity tests; double programming (two model developers program components independently and/or the model is programmed in two different software packages to determine if the same results are obtained).
